# Frailty and comorbidity in predicting community COVID-19 mortality in the UK Biobank: the effect of sampling

**DOI:** 10.1101/2020.10.22.20217489

**Authors:** Jonathan K. L. Mak, Ralf Kuja-Halkola, Yunzhang Wang, Sara Hägg, Juulia Jylhävä

**Author notes:** **Corresponding author** Jonathan K. L. Mak, Department of Medical Epidemiology and Biostatistics, Karolinska Institutet, Nobels väg 12A, 171 65 Stockholm, Sweden.

## Abstract

Frailty has been linked to increased risk of COVID-19 mortality, but evidence is mainly limited to hospitalized older individuals and analyses in community samples are scarce. This study aims to assess and compare the predictive abilities of different frailty measures – the frailty phenotype (FP), frailty index (FI), and Hospital Frailty Risk Score (HFRS), and comorbidity, measured using the Charlson Comorbidity Index (CCI), on COVID-19 mortality in a UK community sample of adults aged 52–86 years. We analyzed (i) the full sample of 428,754 UK Biobank participants and (ii) a subsample of 2,287 COVID-19 positive UK Biobank participants with data on COVID-19 outcomes between March 1 and September 21, 2020. COVID-19 positivity was confirmed by PCR, hospital records and/or death registers. Logistic regression models adjusted for age, sex, smoking, ethnicity, and socioeconomic variables with areas under the receiver operating characteristic curves (AUCs) were used in the modelling. Overall, 391 individuals died of COVID-19. In the full sample, all frailty measures and the CCI were associated with COVID-19 mortality but only the HFRS and CCI improved the predictive ability of a model including age and sex, yielding AUCs>0.80. However, when restricting analyses to the COVID-19 positive subsample, which had an over-representation of frail individuals, similar improvement in AUCs was not observed in which only the CCI was significantly associated with COVID-19 mortality. Our results suggest that HFRS and CCI can be used in COVID-19 mortality risk stratification at the population level, but they show limited added value in COVID-19 positive individuals.

## Introduction

Coronavirus disease 2019 (COVID-19), caused by the severe acute respiratory syndrome coronavirus 2 (SARS-CoV-2), has led to a global pandemic, affecting more than 59 million individuals and causing ∼1.4 million deaths worldwide as of November 24, 2020 [1]. Accumulating evidence has shown that older age, male sex, comorbidities (e.g. diabetes, hypertension), and laboratory indicators (e.g. elevated levels of d-dimer, interleukin 6) are risk factors for COVID-19-associated mortality [2–4]. However, there are relatively few data available for risk stratification in community samples compared to hospitalized patients. With the continuing spread of the SARS-CoV-2 across the world, there is an urgent need to identify the strongest determinants of COVID-19 mortality to target the high-risk groups in the general population and mitigate the impact of COVID-19.

Frailty, characterized as a state of increased vulnerability due to cumulative decline in multiple physiological systems [5], has consistently shown to be a strong predictor of mortality in the general population [6,7]. Various scales have been developed for measuring frailty, some of which require in-person assessment by a trained healthcare professional. One example is the Clinical Frailty Scale (CFS) [8], that is frequently used in clinical settings. Other widely used measures include the Fried frailty phenotype (FP), which defines frailty as a clinical syndrome associated with unintentional weight loss, exhaustion, slowness, low physical activity, and weakness [9]; and the Rockwood frailty index (FI), which is a multidimensional measure of frailty and defined as a ratio of accumulated deficits over the total number of deficits considered [10]. The Hospital Frailty Risk Score (HFRS), constructed based on *International Statistical Classification of Diseases and Related Health Problems, Tenth Revision* (ICD-10) codes [11], was developed for frailty risk stratification among older hospitalized patients and has been validated for its ability to predict adverse outcomes in various hospital settings [12,13].

A growing number of studies have investigated the association between frailty, frequently measured using the CFS, and mortality among COVID-19 patients in hospital settings, most of which suggested that frailty may add to the risk prediction in hospitalized patients [14–17], although some studies observed only weak [18,19], or even null associations [20]. Some inconsistencies in prior results may partly be owing to the COVID-19 patients being a non-random, selected sample with a particularly high prevalence of frailty [21], potentially causing a selection bias [22]. For community samples, a recently published study in the UK Biobank observed that frailty measured using the FP or FI at baseline assessment during 2006–2010 was associated with elevated risks of COVID-19-associated hospitalizations and deaths [23]. However, as these frailty measures were assessed approximately a decade ago before the COVID-19 pandemic, they may not fully reflect one’s current physiological status as frailty can change across the adult lifespan [24]. Moreover, the FP and FI may be of limited use outside research settings as they cannot be constructed from patients’ medical records. Lastly, it has not been studied whether equally accurate predictions can be obtained using easily accessible comorbidity indices, such as the Charlson comorbidity index (CCI). It is thus important to assess whether COVID-19 mortality can be accurately predicted using concurrent frailty and comorbidity measures, such as the HFRS and CCI that can be readily derived from medical records at any given time.

In this study, our overarching goal was to assess whether an easily accessible frailty and/or comorbidity measure could aid in COVID-19 mortality risk stratification and provide added value beyond demographic predictors, such as age and sex in community settings. Using the large population-based UK Biobank cohort, we aimed to investigate and compare the predictive abilities of frailty, measured using the FP, FI, and HFRS, and comorbidity measured using the CCI for COVID-19 mortality in (i) the overall community population and (ii) COVID-19 positive individuals. Such comparisons are currently lacking, yet of importance as selected samples, such as those already being tested positive for COVID-19 are likely to differ in many characteristics from the overall population. Lastly, in keeping with the observations that higher frailty levels carry a relatively greater risk of all-cause mortality in younger than older ages [6,7], we additionally assessed whether the same holds true for COVID-19 mortality. As excess mortality due to COVID-19 may be more pronounced in younger and fitter individuals compared to old and frail [20], it is essential to identify the factors contributing to the risk.

## Methods

### Study population

This is a population-based cohort study using data from the UK Biobank. Between 2006 and 2010, more than 500,000 participants completed a touch-screen questionnaire, had physical measurements taken, and provided biological samples at one of the 22 assessment centers throughout the UK [25]. The UK Biobank study was approved by the North West Multi-Centre Research Ethics Committee. All participants provided written informed consent for data collection, analysis, and record linkage.

We excluded participants who died before March 1, 2020, requested to withdraw from the study prior to August 2020, and had missing data on frailty and comorbidity measures. This resulted in a sample size of *n*=428,754, which we refer to as the “full sample”. The subgroup of “COVID-19 positive subsample” (*n*=2,287) consisted of those who were tested positive, diagnosed as COVID-19 patients in hospitals, and/or died of COVID-19. The analyses were performed analogously in both samples (Supplementary Fig. 1).

### COVID-19 diagnosis and mortality

Information on COVID-19 was obtained from three data sources linked to the UK Biobank: laboratory test results, inpatient medical records, and death register. SARS-CoV-2 polymerase chain reaction (PCR) test results were provided by Public Health England [26], with data available in England only, between March 16 and September 21, 2020. Hospital inpatient data were sourced from the Hospital Episode Statistics (HES), containing electronic medical records (i.e., ICD-10 codes) for all hospital admissions to National Health Service (NHS) hospitals in England up to June 30, 2020. Death register data included all deaths up until September 21, 2020 in England, Wales and Scotland, containing ICD-10 codes assigned as individuals’ primary and contributory causes of death.

Participants were considered as “COVID-19 positive” when meeting at least one of the following criteria: (i) being positive in at least one of the PCR tests; (ii) shown as COVID-19 inpatients, with ICD-10 code U07 in hospital admission; and (iii) died of COVID-19, defined as those with COVID-19 (ICD-10 code U07) as the primary or contributory causes of death. COVID-19 mortality was used as the main outcome in the analyses.

### Frailty and comorbidity measures

Frailty was assessed using the FP, FI, and HFRS, and comorbidity was measured using the CCI. A modified FP has previously been created by Hanlon *et al*. for UK Biobank participants [27], based on the five frailty criteria in the original Fried FP [9]. Weight loss, exhaustion, slowness, and low physical activity were self-reported from baseline questionnaire, while weakness was assessed by measured grip strength at baseline, where the higher value of the right- and left-hand measurements were used in analysis (Supplementary Table 1). The FI has previously been created and validated by us for UK Biobank participants, using 49 self-reported frailty items assessed at baseline during 2006–2010 that cover a wide range of items for physical and mental well-being (Supplementary Table 2) [7]. The FI was calculated as the sum of items (deficits) present in an individual divided by the total (e.g., an individual with 7 deficits of the 49 would receive an FI of 7/49=0.14). The FI was categorized into relatively fit (≤0.03), less fit (>0.03–0.1), least fit (>0.1–0.21) and frail (>0.21) [24]. The HFRS and CCI were computed based on ICD-10 codes from hospital records [11,28]. Only medical records in the prior 2 years (i.e., between March 1, 2018 and February 29, 2020) were included for calculation. As previously described by Gilbert *et al*., each of the 109 frailty-related ICD-10 codes were assigned a weight ranging from 0.1 to 7.1, depending on its strength of the association with frailty (Supplementary Table 3) [11]. The HFRS was then calculated by summing all the weighted codes, and categorized into low (<5), intermediate (5–15) and high (>15) risk of frailty [11]. Similarly, the CCI was derived by summing weighted ICD-10 codes of 17 comorbidities, with weights ranging from 1 to 6 depending on disease severity and mortality risk (Supplementary Table 4) [28], and was treated as a continuous variable in all analyses. Individuals who had missing hospital data were those who had not been hospitalized or resided outside England (these data were only available for England). To maximize data utilization, we first excluded individuals who attended baseline assessment in Wales or Scotland and with missing hospital data; and then for the remaining individuals with missing hospital data, who were likely to be younger and healthier individuals and never been hospitalized (Supplementary Table 5), we coded them as 0 for HFRS and CCI.

### Other study variables

Demographic characteristics and socioeconomic indicators were collected at baseline during 2006–2010. Education was assessed by the highest self-reported qualification and categorized into low (no relevant qualifications), intermediate (A levels, O levels/GCSEs, CSEs, NVQ/HND/HNC, other professional qualifications) and high (college or university degree). Annual household income was self-reported and categorized into four groups (<£18,000, £18,000–30,999, £31,000–51,999, ≥£52,000). Townsend deprivation index was derived from national census data regarding unemployment, car ownership, home ownership, and household overcrowding; higher scores correspond to higher levels of socioeconomic deprivation.

### Statistical analyses

Descriptive statistics were calculated for the full sample and COVID-19 positive subsample. Study variables were compared between those who were classified in the COVID-19 positive subsample and those who were not, using t-tests or Mann–Whitney *U* tests for continuous variables, and χ^2^-tests for categorical variables. Due to the apparent over-representation of frail individuals in the COVID-19 positive subsample, we performed logistic regression to formally ascertain if frailty and comorbidity were determinants for being COVID-19 positive.

In both samples, odds ratios (ORs) with 95% confidence intervals (CI), in multivariable logistic regression models were used to investigate the associations of frailty and comorbidity (FP, FI, HFRS, and CCI) with COVID-19 mortality, adjusted for age (as linear effect, after confirming that the age-mortality relationship was approximately linear) and sex. Ethnicity, smoking status, and socioeconomic variables were subsequently added into the models to test whether they had an effect on the associations. Areas under the receiver operating characteristic curves (AUC) were used to assess the predictive accuracies of the different measures. Because the HFRS was originally designed for individuals older than 75 years and previous studies have reported age-varying risks for frailty [6,7], we stratified the analysis by age (<65, 65–74, and ≥75 years), as well as performed an analysis with an interaction term between HFRS and age as continuous variables. With these approaches, we aimed to discern whether the risk carried by higher frailty is age-varying. As a sensitivity analyses to assess the robustness of our findings, we performed multinomial logistic regression models to account for non-COVID-19 deaths as competing risk, where mortality due to COVID-19 or other causes than COVID-19 were compared to those who were alive as of September 21, 2020.

Spearman’s correlations were calculated between frailty and comorbidity measures, and between socioeconomic variables to determine whether they can be included in the same model. Multicollinearity was inspected in all regression models using variance inflation factors. To account for multiple comparisons, the Benjamini-Hochberg false discovery rate method was applied [29]. All analyses were performed using Stata v16.0 (Stata Corp, College Station, TX) and R v3.6.3 (R Foundation for Statistical Computing, Vienna, Austria).

## Results

### Sample characteristics

In the full sample of 428,754 participants, the mean age as of January 2020 was 68.1 (standard deviation [SD] 8.1) and 55.2% were women (**Table 1**). 2,287 individuals from the full sample were considered as COVID-19 positive, who were either diagnosed as COVID-19 patients (i.e., had positive PCR results or were COVID-19 inpatients; *n*=2,201), or died of COVID-19 but without positive test record (*n*=86); both groups were generally comparable except that individuals in the latter group were more likely to be older, with lower income and with higher HFRS (Supplementary Table 6). When comparing individuals who were classified as COVID-19 positive than those who were not, we observed significantly greater proportions of the oldest age group, men, Black ethnicity, previous or current smokers, low education, lowest income and most deprived groups in the COVID-19 positive subsample (all *p*<0.001). The different frailty (FP, FI, and HFRS) and comorbidity (CCI) measures had small-to-moderate correlations with each other (Spearman’s correlations 0.12–0.55) (Supplementary Table 7). Frailty appeared to be over-represented among COVID-19 positive individuals, with 8.1%, 19.6% and 11.9% in the most frail group as assessed by the FP, FI and HFRS, respectively, compared to that of 3.4%, 11.9%, and 0.9% in the full sample. Logistic regression models also showed that the FP, FI, HFRS, and CCI were all associated with higher risk of being COVID-19 positive, after adjusting for age and sex (Supplementary Table 8).

**Table 1.** Characteristics of UK Biobank participants in the full sample and COVID-19 positive subsample

| Characteristic | Full sample<br>(n=428,754) | Classified as COVID-19 positive<sup>a</sup> |  | *p*<sup>b</sup> |
| --- | --- | --- | --- | --- |
|  |  | No (n=426,467) | Yes (n=2,287) |  |
| Deaths, n (%) | 2,351 (0.6) | 1,882 (0.4) | 469 (20.5) | <0.001 |
| Age (year), mean (SD) | 68.1 (8.1) | 68.1 (8.1) | 68.1 (9.0) | 0.75 |
| Age category, n (%) |  |  |  | <0.001 |
| <65 | 145,300 (33.9) | 144,437 (33.9) | 863 (37.7) |  |
| 65–74 | 173,507 (40.5) | 172,842 (40.5) | 665 (29.1) |  |
| ≥75 | 109,947 (25.6) | 109,188 (25.6) | 759 (33.2) |  |
| Sex, n (%) |  |  |  | <0.001 |
| Female | 236,644 (55.2) | 235,547 (55.2) | 1,097 (48.0) |  |
| Male | 192,110 (44.8) | 190,920 (44.8) | 1,190 (52.0) |  |
| Ethnicity, n (%) |  |  |  | <0.001 |
| White | 403,410 (94.4) | 401,403 (94.4) | 2,007 (88.1) |  |
| Asian | 9,906 (2.3) | 9,794 (2.3) | 112 (4.9) |  |
| Black | 7,351 (1.7) | 7,247 (1.7) | 104 (4.6) |  |
| Others | 6,641 (1.6) | 6,585 (1.6) | 56 (2.5) |  |
| Smoking status, n (%) |  |  |  | <0.001 |
| Never | 237,707 (55.6) | 236,603 (55.7) | 1,104 (48.6) |  |
| Previous | 147,477 (34.5) | 146,590 (34.5) | 887 (39.1) |  |
| Current | 42,111 (9.9) | 41,831 (9.8) | 280 (12.3) |  |
| Education, n (%) |  |  |  | <0.001 |
| Low | 68,631 (16.2) | 68,078 (16.1) | 553 (24.6) |  |
| Intermediate | 216,335 (51.0) | 215,199 (51.0) | 1,136 (50.4) |  |
| High | 139,621 (32.9) | 139,057 (32.9) | 564 (25.0) |  |
| Income, n (%) |  |  |  | <0.001 |
| <£18,000 | 79,917 (21.7) | 79,306 (21.7) | 611 (31.9) |  |
| £18,000–30,999 | 93,403 (25.4) | 92,919 (25.4) | 484 (25.3) |  |
| £31,000–51,999 | 97,403 (26.5) | 96,972 (26.5) | 431 (22.5) |  |
| ≥£52,000 | 96,962 (26.4) | 96,571 (26.4) | 391 (20.9) |  |
| Townsend deprivation quintile, n (%) |  |  |  | <0.001 |
| 1 (least deprived) | 86,444 (20.2) | 86,101 (20.2) | 343 (15.0) |  |
| 2 | 87,408 (20.4) | 87,029 (20.4) | 379 (16.6) |  |
| 3 | 86,805 (20.3) | 86,389 (20.3) | 416 (18.2) |  |
| 4 | 85,812 (20.0) | 85,329 (20.0) | 483 (21.1) |  |
| 5 (most deprived) | 81,773 (19.1) | 81,108 (19.0) | 665 (29.1) |  |
| Frailty phenotype category, n (%) |  |  |  | <0.001 |
| Non-frail | 248,820 (58.1) | 247,689 (58.1) | 1,131 (49.5) |  |
| Pre-frail | 165,192 (38.5) | 164,220 (38.5) | 972 (42.5) |  |
| Frail | 14,742 (3.4) | 14,558 (3.4) | 184 (8.1) |  |
| Frailty index category, n (%) |  |  |  | <0.001 |
| Relatively fit | 25,898 (6.0) | 25,800 (6.1) | 98 (4.3) |  |
| Less fit | 162,149 (37.8) | 161,451 (37.9) | 698 (30.5) |  |
| Least fit | 189,736 (44.3) | 188,694 (44.3) | 1,042 (45.6) |  |
| Frail | 50,971 (11.9) | 50,522 (11.9) | 449 (19.6) |  |
| Hospital Frailty Risk Score category, n (%) |  |  |  | <0.001 |
| Low risk | 409,348 (95.5) | 407,563 (95.6) | 1,785 (78.1) |  |
| Intermediate risk | 15,599 (3.6) | 15,369 (3.6) | 230 (10.1) |  |
| High risk | 3,807 (0.9) | 3,535 (0.8) | 272 (11.9) |  |
| Charlson Comorbidity Index score, mean (SD) | 0.28 (0.88) | 0.28 (0.87) | 0.94 (1.72) | <0.001 |

<sup>a</sup> Individuals were considered as COVID-19 positive if (i) being positive in at least one of the PCR tests, (ii) was a COVID-19 inpatient as shown in hospital records, and/or (iii) died of COVID-19
<sup>b</sup> Based on t-tests or Mann–Whitney *U* tests for continuous variables and $\chi^2$ -tests for categorical variables when comparing the COVID-19 positive vs. non-COVID-19 positive subgroups
Abbreviations: *IQR*, interquartile range; *SD*, standard deviation

### Frailty and comorbidity in predicting COVID-19 mortality

In total, 391 individuals died of COVID-19 between March 1 and September 21, 2020. Results for the logistic regression models of the different frailty measures and CCI on COVID-19 mortality are presented in **Table 2**. In the full sample, when each of the measures – FP, FI, HFRS, and CCI were tested separately, they were significantly associated with higher odds of COVID-19 mortality after controlling for age and sex (models 2–5). The AUC for the model including only age and sex was 0.76 (95% CI 0.74–0.78); adding FP and FI to the model resulted in very small, yet statistically significantly improved AUCs of 0.77 (95% CI 0.75–0.80) and 0.78 (95% CI 0.75–0.80), respectively (*p*<0.001 for pairwise comparisons). Meanwhile, adding HFRS and CCI resulted in even greater improvement in AUCs, both yielding an AUC of 0.82 (95% CI 0.80–0.84) (*p*<0.001 for pairwise comparisons) (**Fig. 1A**). When adding all the frailty measures and CCI to the same model (model 6 in **Table 2 panel a**), the CCI (OR 1.19; 95% CI 1.13–1.26) and the HFRS categories of intermediate and high risk (OR for high vs. low risk=11.21; 95% CI 8.06–15.57) predicted a higher risk of COVID-19 mortality, whereas the association of the FP was attenuated so that only the frail category remained significantly associated with mortality while the pre-frail category did not. None of the FI categories (less fit, least fit and frail) were significantly associated with mortality in this model.

**Table 2.** Associations between different frailty and comorbidity measures, and COVID-19 mortality in the full sample and COVID-19 positive subsample

| Variable | Model 1<sup>a</sup> | Model 2<sup>a</sup> | Model 3<sup>a</sup> | Model 4<sup>a</sup> | Model 5<sup>a</sup> | Model 6<sup>a</sup> |
| --- | --- | --- | --- | --- | --- | --- |
|  | OR (95% CI) | OR (95% CI) | OR (95% CI) | OR (95% CI) | OR (95% CI) | OR (95% CI) |
| **(a) Full sample (n=428,754)** |  |  |  |  |  |  |
| Age | 1.14 (1.12–1.16)* | 1.14 (1.12–1.16)* | 1.14 (1.12–1.16)* | 1.11 (1.09–1.13)* | 1.12 (1.10–1.14)* | 1.11 (1.08–1.13)* |
| Male sex | 2.04 (1.66–2.51)* | 2.15 (1.75–2.65)* | 2.14 (1.74–2.64)* | 1.96 (1.59–2.41)* | 1.79 (1.46–2.21)* | 1.92 (1.56–2.37)* |
| FP category (ref. non-frail) |  |  |  |  |  |  |
| Pre-frail | - | 1.55 (1.25–1.91)* | - | - | - | 1.19 (0.95–1.48) |
| Frail | - | 3.76 (2.65–5.34)* | - | - | - | 1.54 (1.04–2.29)* |
| FI category (ref. relatively fit) |  |  |  |  |  |  |
| Less fit | - | - | 1.41 (0.74–2.71) | - | - | 1.33 (0.69–2.55) |
| Least fit | - | - | 2.00 (1.06–3.78)* | - | - | 1.51 (0.80–2.87) |
| Frail | - | - | 3.72 (1.94–7.15)* | - | - | 1.67 (0.85–3.29) |
| HFRS category (ref. low risk) |  |  |  |  |  |  |
| Intermediate | - | - | - | 5.24 (4.00–6.87)* | - | 3.42 (2.53–4.62)* |
| High risk | - | - | - | 21.81 (16.81–28.30)* | - | 11.21 (8.06–15.57)* |
| CCI score | - | - | - | - | 1.51 (1.45–1.57)* | 1.19 (1.13–1.26)* |
| AUC (95% CI) | 0.76 (0.74–0.78) | 0.77 (0.75–0.80) | 0.78 (0.75–0.80) | 0.82 (0.80–0.84) | 0.82 (0.80–0.84) | 0.84 (0.82–0.86) |
| **(b) COVID-19 positive sample<sup>b</sup> (n=2,287)** |  |  |  |  |  |  |
| Age | 1.13 (1.11–1.15)* | 1.13 (1.11–1.15)* | 1.13 (1.11–1.15)* | 1.12 (1.10–1.14)* | 1.12 (1.10–1.14)* | 1.12 (1.10–1.14)* |
| Male sex | 1.47 (1.16–1.87)* | 1.50 (1.18–1.91)* | 1.49 (1.18–1.90)* | 1.48 (1.17–1.89)* | 1.42 (1.12–1.81)* | 1.45 (1.14–1.85)* |
| FP category (ref. non-frail) |  |  |  |  |  |  |
| Pre-frail | - | 1.18 (0.92–1.50) | - | - | - | 1.12 (0.86–1.45) |
| Frail | - | 1.25 (0.83–1.89) | - | - | - | 1.04 (0.66–1.64) |
| FI category (ref. relatively fit) |  |  |  |  |  |  |
| Less fit | - | - | 1.13 (0.54–2.34) | - | - | 1.10 (0.53–2.30) |
| Least fit | - | - | 1.23 (0.60–2.51) | - | - | 1.14 (0.55–2.33) |
| Frail | - | - | 1.37 (0.66–2.86) | - | - | 1.12 (0.52–2.37) |
| HFRS category (ref. low risk) |  |  |  |  |  |  |
| Intermediate risk | - | - | - | 1.63 (1.17–2.27)* | - | 1.28 (0.89–1.83) |
| High risk | - | - | - | 1.32 (0.97–1.81) | - | 0.94 (0.65–1.35) |
| CCI score | - | - | - | - | 1.14 (1.08–1.21)* | 1.13 (1.06–1.22)* |
| AUC (95% CI) | 0.76 (0.73–0.78) | 0.76 (0.74–0.78) | 0.76 (0.74–0.78) | 0.76 (0.74–0.78) | 0.77 (0.74–0.79) | 0.77 (0.74–0.79) |

<sup>a</sup> Model 1: age (continuous) and sex; models 2–5: added FP, FI, HFRS, and CCI as independent variables respectively; model 6: all listed variables were mutually adjusted for
<sup>b</sup> The COVID-19 positive subsample is nested within the full sample
\* Significant with a false discovery rate corrected significance level at 0.034
Abbreviations: *AUC*, area under the receiver operating characteristic curves; *CCI*, Charlson Comorbidity Index; *CI*, confidence interval; *FI*, frailty index; *FP*, frailty phenotype; *HFRS*, Hospital Frailty Risk Score; *OR*, odds ratio.

**Table 3.** Associations between the Hospital Frailty Risk Score and COVID-19 mortality adjusted for potential confounders in the full sample and COVID-19 positive subsample

| Variable<sup>a</sup> | Full sample (n=428,754) | COVID-19 positive subsample<sup>b</sup> (n=2,287) |
| --- | --- | --- |
|  | OR (95% CI) | OR (95% CI) |
| Age | 1.11 (1.09–1.13)* | 1.14 (1.11–1.16)* |
| Male sex | 1.95 (1.53–2.49)* | 1.49 (1.12–1.98)* |
| HFRS category (ref. low risk) |  |  |
| Intermediate risk | 3.30 (2.34–4.65)* | 1.20 (0.80–1.82) |
| High risk | 11.73 (8.12–16.96)* | 1.01 (0.66–1.54) |
| CCI score | 1.18 (1.10–1.25)* | 1.13 (1.04–1.22)* |
| Ethnicity (ref. white) |  |  |
| Asian | 1.40 (0.65–3.02) | 0.88 (0.38–2.05) |
| Black | 4.70 (2.75–8.04)* | 2.68 (1.37–5.24)* |
| Others | 1.40 (0.52–3.81) | 0.85 (0.28–2.54) |
| Smoking status (ref. never) |  |  |
| Previous | 1.36 (1.06–1.75)* | 1.10 (0.82–1.49) |
| Current | 1.58 (1.10–2.26)* | 1.39 (0.91–2.13) |
| Education (ref. high) |  |  |
| Intermediate | 1.10 (0.82–1.48) | 0.91 (0.64–1.30) |
| Low | 1.23 (0.87–1.74) | 0.84 (0.56–1.26) |
| Income (ref. >£52,000) |  |  |
| £31,000–51,999 | 1.18 (0.77–1.82) | 1.32 (0.81–2.16) |
| £18,000–30,999 | 1.08 (0.70–1.65) | 0.93 (0.57–1.53) |
| <£18,000 | 1.49 (0.97–2.28) | 1.16 (0.70–1.92) |
| Townsend deprivation quintile (ref. 1) |  |  |
| 2 | 0.77 (0.52–1.15) | 0.74 (0.47–1.19) |
| 3 | 0.81 (0.55–1.21) | 0.71 (0.45–1.13) |
| 4 | 1.08 (0.75–1.56) | 0.90 (0.58–1.40) |
| 5 (most deprived) | 1.42 (0.99–2.03) | 1.15 (0.74–1.78) |
| AUC (95% CI) | 0.86 (0.84–0.88) | 0.79 (0.77–0.82) |

<sup>a</sup> All listed variables were mutually adjusted for in the models
<sup>b</sup> The COVID-19 positive subsample is nested within the full sample
\* Significant with a false discovery rate corrected significance level at 0.016
Abbreviations: *AUC*, area under the receiver operating characteristic curves; *CCI*, Charlson Comorbidity Index; *CI*, confidence interval; *HFRS*, Hospital Frailty Risk Score; *OR*, odds ratio

**Fig. 1.**
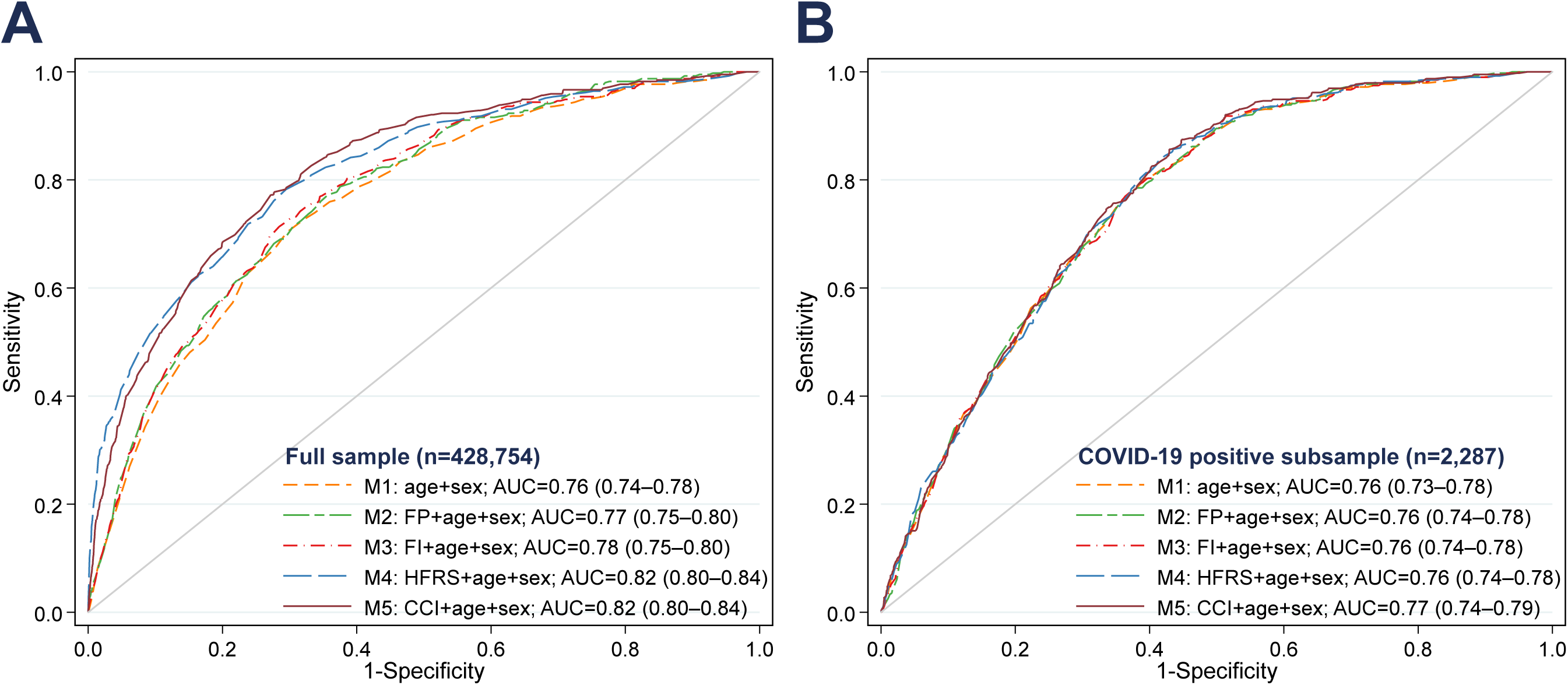
Receiver operating characteristic curves for age, sex, frailty and comorbidity in predicting COVID-19 mortality in the full sample and COVID-19 positive subsample. The COVID-19 positive subsample is nested within the full sample. Model 1: age and sex; models 2–5: added FP, FI, HFRS, and CCI as independent variables respectively. Pairwise comparisons of AUCs in the full sample: *p*<0.001 for models 2–5 vs. model 1. Pairwise comparisons of AUCs in the COVID-19 positive sample: *p*=0.43 for model 2 vs. model 1; *p*=0.62 for model 3 vs. model 1; *p*=0.19 for model 4 vs. model 1; *p*=0.07 for model 5 vs. model 1. Abbreviations: *AUC*, area under the receiver operating characteristic curves; *CCI*, Charlson Comorbidity Index; *FI*, frailty index; *FP*, frailty phenotype; *HFRS*, Hospital Frailty Risk Score.

After restricting the sample to COVID-19 positive individuals, all of these associations were attenuated, and the predictive accuracies decreased; only the CCI, but none of the frailty measures remained significantly associated with COVID-19 mortality in a model including all the frailty measures (model 6 in **Table 2 panel b**). In this sample, frailty and comorbidity did not add predictive value on top of age and sex, as indicated by similar AUCs across all models (all *p*>0.05 for pairwise comparisons) (**Fig. 1B**).

To further assess whether the predictiveness of HFRS for COVID-19 mortality would change after adjusting for potential confounders, we subsequently adjusted for ethnicity, smoking and socioeconomic variables, and observed that the associations and AUCs remained essentially unchanged in both samples (**Table 3**).

Compared with the oldest old individuals (≥75 years), relatively younger individuals (<75 years) had higher ORs for COVID-19 mortality across HFRS categories in both samples (**Fig. 2**). There was also a significant interaction between HFRS and age in both the full sample (*P*_interaction_=0.003) and COVID-19 positive subsample (*P*_interaction_<0.001), so that higher frailty carried a relatively greater risk in younger adults (Supplementary Table 9). We also tested the interaction between HFRS and sex, but it was not statistically significant, and thus we did not perform further analysis stratified by sex.

**Fig. 2.**
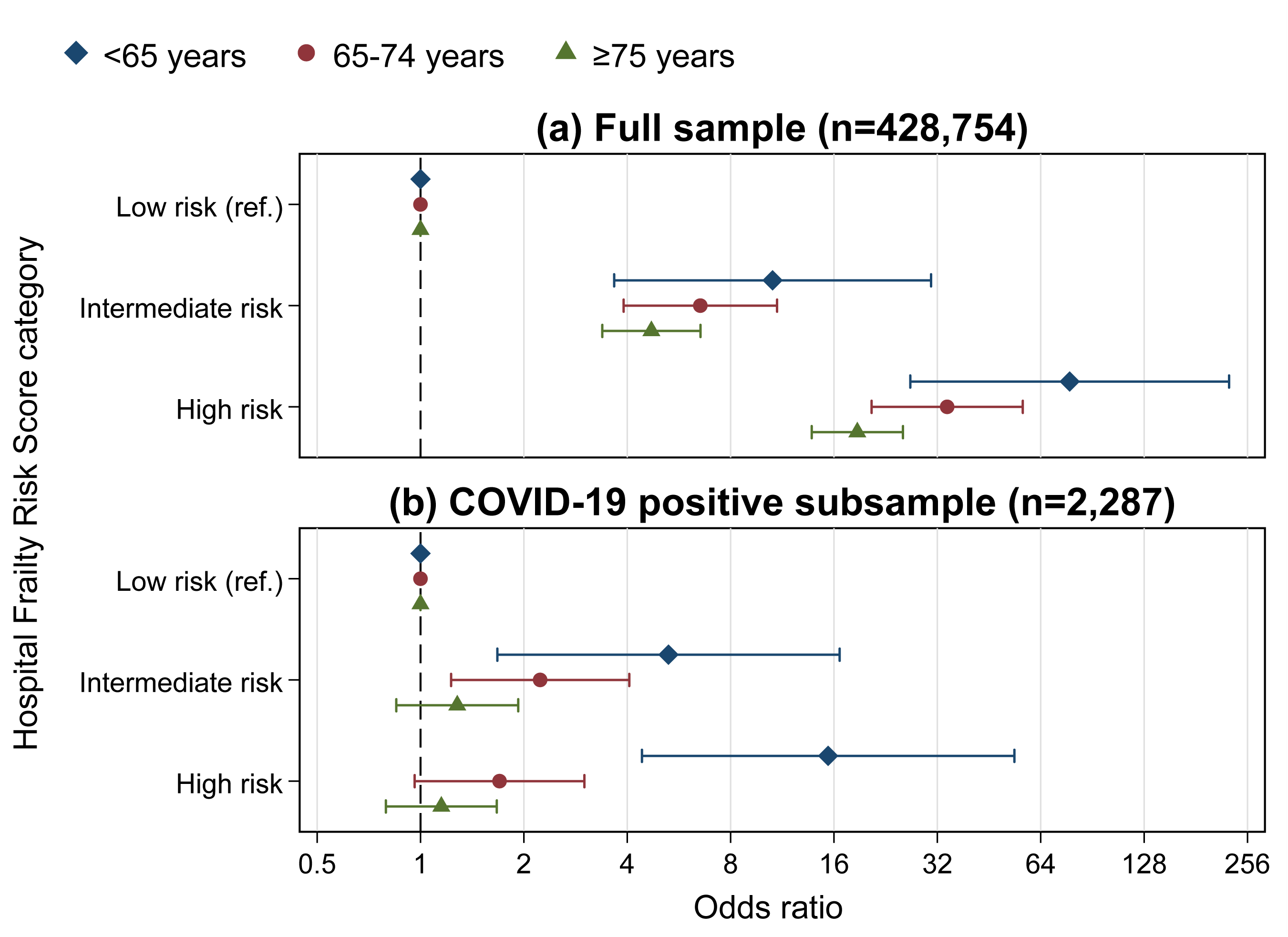
Associations between the Hospital Frailty Risk Score and COVID-19 mortality stratified by age in the full sample and COVID-19 positive subsample. The COVID-19 positive subsample is nested within the full sample. Error bars indicate 95% confidence intervals. Models were adjusted for sex. Point estimates for the stratified analysis can be found in the Supplementary Table 9.

### Sensitivity analyses

The associations between the frailty measures and CCI with COVID-19 mortality were largely similar as in the main analyses when accounting for competing risk by deaths due to other causes than COVID-19 (Supplementary Table 10).

## Discussion

Using data from the UK Biobank, we found that the HFRS and CCI, measures of frailty and comorbidity, respectively, constructed using the ICD-10 codes from medical records, were viable and independent predictors of COVID-19 mortality and added predictive value on top of age and sex in the overall community population. Equally accurate predictions could be obtained by including either the HFRS or CCI to a model with age and sex, in which addition of either one resulted in an improvement of AUC from 0.76 to 0.82. The associations persisted even after adjusting for ethnicity, smoking and socioeconomic variables. However, in the COVID-19 positive subsample that had an over-representation of frail individuals, neither the frailty measures nor the CCI improved the predictive accuracy of a model including age and sex. Stronger associations between HFRS and COVID-19 mortality were seen among younger old (<75 years) than older old individuals (≥75 years), indicating that the HFRS may be applicable for predicting mortality risk in younger adults as well.

To the best of our knowledge, this is the first study that has utilized the HFRS as a frailty measure for COVID-19 mortality prediction in the community population. While it has previously been shown that a FP or FI was associated with higher risk of COVID-19 mortality in the full sample of the UK Biobank [23], both measures were assessed at the UK Biobank baseline ∼10 years ago, which may affect their predictive accuracies. By way of how the FP and FI are constructed (in-person testing and a 49-item questionnaire, respectively), they may also be less feasible in clinical decision-making or to be implemented in population risk stratification during the current COVID-19 pandemic. Adding to the existing literature, we found that the HFRS constructed based on ICD-10 codes from the past 2 years was a stronger predictor than the FP or FI for COVID-19 mortality. The CCI, a measure of comorbidity computed by ICD-10 codes, likewise predicted COVID-19 mortality, which is in line with prior research showing a positive association between comorbidity and COVID-19 deaths [16,19]. Our results thus suggest that frailty and comorbidity measures available in routinely collected medical records may be used in identification of at-risk individuals in the community.

However, the predictive accuracies of frailty and comorbidity for COVID-19 mortality reduced after restricting the sample to those infected. It has been argued that using non-random samples may induce selection bias in COVID-19-related studies [22]. In our COVID-19 positive subsample, we found an over-representation of the most frail individuals. Moreover, consistent with a previous study, we confirmed that frailty and comorbidities are independent determinants of COVID-19 positivity [30]. Such an over-representation of frailty may partly explain the inconsistencies in the frailty-mortality associations among hospitalized COVID-19 patients [18–20]. In a relatively frail sample, such as the COVID-19 positive group in our study, the mortality risk may be more related to other factors, such as viral load and host immune characteristics [18,31]. Indeed, none of our models in the COVID-19 positive subsample yielded a good predictive accuracy of AUC>0.8, even when smoking, ethnicity and socioeconomic variables were included. More research is thus warranted to identify the most accurate predictors for mortality among COVID-19 patients.

Given that the HFRS was initially developed for older individuals, we stratified the analysis by age and observed a more pronounced association between HFRS and COVID-19 mortality among individuals younger than 75 years old. A similar pattern has also been reported previously, with higher frailty being more strongly associated with all-cause and cause-specific mortality at midlife than in the oldest ages [6]. These findings indicate that although the absolute risk of death is always higher with advancing age, frailty carries a relatively greater risk at younger ages. Our present findings thus highlight the importance of frailty screening in younger individuals in prevention for COVID-19-related mortality.

The large sample of UK Biobank participants with linkage to COVID-19 data enabled us to study the associations among the overall population and to examine the potential effects of sample selection. Nevertheless, there are several limitations to this study. Firstly, the HFRS was originally designed for hospitalized individuals rather than for the general population, although we attempted to account for individuals with missing hospital data and showed that it may be applied to community samples as well. Alternatively, future research may utilize frailty measures based on routine primary care data, such as the electronic FI [32] for assessing its predictive ability for COVID-19 mortality. Secondly, during the earlier periods of the epidemic, COVID-19 testing in the UK was largely restricted to hospitalized individuals, who have more severe course of the disease. As such, mild or asymptomatic COVID-19 cases may conceivably be missed, leading to an underestimation of COVID-19 positive cases. Thirdly, we modelled the outcome, COVID-19 mortality, as a binary trait rather than as time-to-event outcome because we could not ascertain the exact date of confirmed COVID-19 infection for several individuals in the COVID-19 positive subsample. However, as the follow-up time was limited, it could be considered essentially complete for most participants (i.e., minimal censoring due to migration and other deaths). Finally, UK Biobank is not a nationally representative sample, with generally healthier and less socioeconomically deprived participants than the UK average [33], thereby reducing the generalizability of our findings.

In conclusion, HFRS and CCI, measures of frailty and comorbidity that can be constructed using routinely collected medical records, predicted COVID-19 mortality in the overall community sample and improved predictive accuracy on top of age and sex, with similar added values with the addition of either one of these measures. Nevertheless, similar effects of added value were not seen in those who already have the disease. Together, our results suggest that identification of frail individuals in the general population may be a viable strategy for COVID-19 mortality risk stratification.

## Data Availability

UK Biobank is an open access resource. All bona fide researchers can apply to use its data for health-related research that is in the public interest (http://www.ukbiobank.ac.uk/register-apply).

## Acknowledgements

This research was conducted using the UK Biobank resource, as part of the registered project 22224.

## Declarations

### Ethical approval

Ethical approval for this study is covered by the general ethics review for the UK Biobank, conducted by North West Multi-Centre Research Ethics Committee (Reference: 16/NW/0274, 13 May 2016).

### Consent to participate

Informed consent was obtained from all individual participants included in the study.

### Consent to publish

Not applicable.

### Code availability

Not applicable.

### Authors’ contributions

JKLM, RK-H, and JJ contributed to the study concept and design; YW, SH and JJ were responsible for acquisition of data; JKLM, RK-H and JJ analyzed and interpreted the data; JKLM and JJ drafted the manuscript; RK-H, YW and SH critically revised the manuscript for important intellectual content. All authors read and approved the final manuscript.

