## Supplementary for "Frailty and comorbidity in predicting community COVID-19 mortality in the UK Biobank: the effect of sampling"

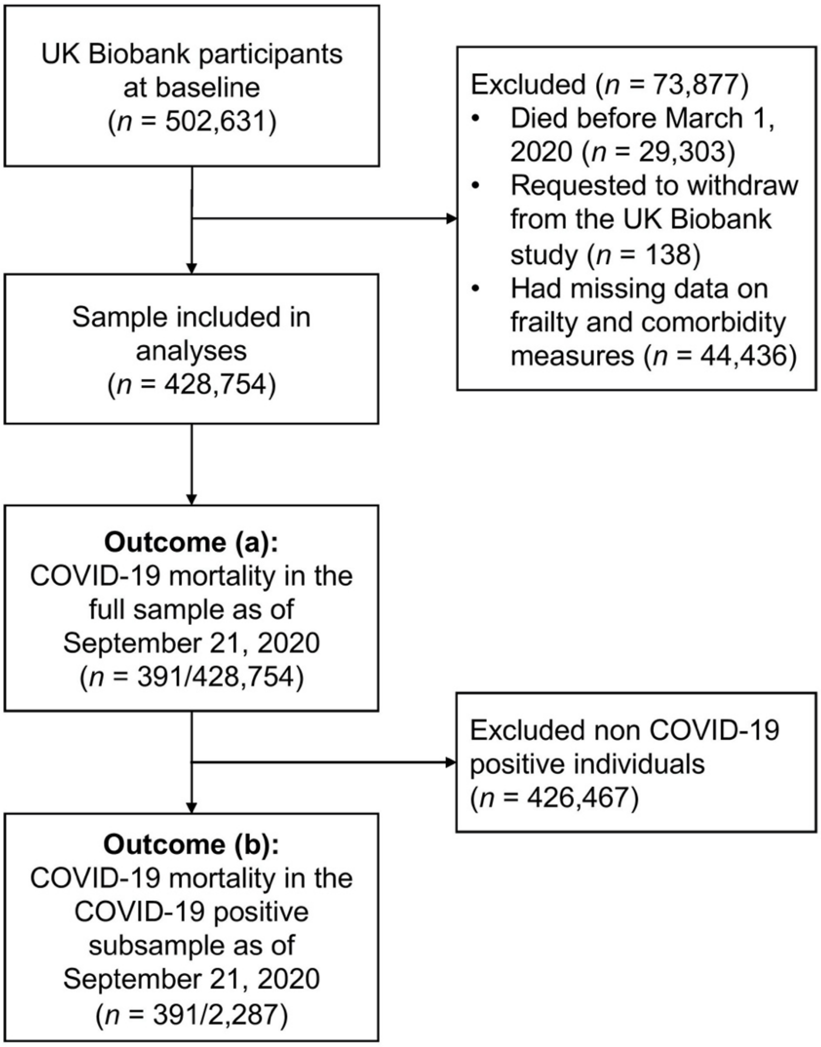

**Supplementary Fig. 1** Flowchart of selection of study samples from the UK Biobank

**Supplementary Table 1** List of the five frailty criteria for construction of the frailty phenotype

| Frailty criteria | Measurement | Scoring |
|---|---|---|
| Weight loss | Self-reported question:<br>"Compared with one year ago, has your weight changed?" | Yes, lost weight=1; other=0 |
| Exhaustion | Self-reported question:<br>"Over the past two weeks, how often have you felt tired or had little energy?" | More than half the days or nearly every day=1; other=0 |
| Slowness | Self-reported question:<br>"How would you describe your usual walking pace?" | Slow=1; other=0 |
| Low physical activity | Self-reported physical activity questionnaire, categorized into 4 levels:<br>(i) none – no physical activity in the last 4 weeks<br>(ii) low – light DIY activity (e.g., pruning, watering the lawn) only in the last 4 weeks<br>(iii) medium – heavy DIY activity (e.g., weeding, lawn mowing, carpentry and digging), walking for pleasure, or other exercises in the last 4 weeks<br>(iv) high – strenuous sports in the last 4 weeks | None or light activity with a frequency of once per week or less=1; medium or heavy activity, or light activity more than once per week=0 |
| Weakness | Measured grip strength | Sex and body mass index (BMI) adjusted cutoffs:<br>Men:<br>≤29 kg for BMI ≤24;<br>≤30 kg for BMI 24.1–26;<br>≤30 kg for BMI 26.1–28; or<br>≤32 kg for BMI >28<br>Women:<br>≤17 kg for BMI ≤23;<br>≤17.3 kg for BMI 23.1–26;<br>≤18 kg for BMI 26.1–29; or<br>≤21 kg for BMI>29 |

**Supplementary Table 2** List of the 49 frailty items and scoring for construction of the frailty index

| # | Frailty item | Scoring |
|---|---|---|
| 1 | Glaucoma | No=0; Yes=1 |
| 2 | Cataracts | No=0; Yes=1 |
| 3 | Hearing difficulty | No=0; Yes/completely deaf=1 |
| 4 | Migraine | No=0; Yes=1 |
| 5 | Dental problems (ulcers, painful gums, bleeding gums, loose teeth, toothache, dentures) | None=0; Any=1 |
| 6 | Self-rated health | Excellent=0; Good=0.25; Fair=0.5; Poor=1 |
| 7 | Fatigue: frequency of tiredness/lethargy in last two weeks | Not at all=0; Several days=0.25; More than half=0.5; Nearly every day=1 |
| 8 | Sleep: experience of sleeplessness/insomnia | Never/rarely=0; Sometimes=0.5; Usually=1 |
| 9 | Depressed feelings: frequency in last two weeks | Not at all=0; Several days=0.5; More than half=0.75; Nearly every day=1 |
| 10 | Self-described nervous personality | No=0; Yes=1 |
| 11 | Severe anxiety/ panic attacks | No=0; Yes=1 |
| 12 | Common to feel loneliness | No=0; Yes=1 |
| 13 | Sense of misery (ever/never) | No=0; Yes=1 |
| 14 | Infirmity: long-standing illness or disability | No=0; Yes=1 |
| 15 | Falls in last year | No falls=0; one fall=0.5; more than one=1 |
| 16 | Fractures/broken bones in last five years | No=0; Yes=1 |
| 17 | Diabetes | No=0; Yes=1 |
| 18 | Myocardial infarction | No=0; Yes=1 |
| 19 | Angina | No=0; Yes=1 |
| 20 | Stroke | No=0; Yes=1 |
| 21 | High blood pressure | No=0; Yes=1 |
| 22 | Hypothyroidism | No=0; Yes=1 |
| 23 | Deep-vein thrombosis | No=0; Yes=1 |
| 24 | High cholesterol | No=0; Yes=1 |
| 25 | Breathing: wheeze in last year | No=0; Yes=1 |
| 26 | Pneumonia | No=0; Yes=1 |
| 27 | Chronic bronchitis/emphysema | No=0; Yes=1 |
| 28 | Asthma | No=0; Yes=1 |
| 29 | Rheumatoid arthritis | No=0; Yes=1 |
| 30 | Osteoarthritis | No=0; Yes=1 |
| 31 | Gout | No=0; Yes=1 |
| 32 | Osteoporosis | No=0; Yes=1 |
| 33 | Hay fever, allergic rhinitis or eczema | No=0; Yes=1 |
| 34 | Psoriasis | No=0; Yes=1 |
| 35 | Any cancer diagnosis | No=0; Yes=1 |
| 36 | Multiple cancers diagnosed | No/single cancer=0; Multiple cancers=1 |
| 37 | Chest pain | No=0; Yes=1 |
| 38 | Head and/or neck pain | No=0; Yes=1 |
| 39 | Back pain | No=0; Yes=1 |
| 40 | Stomach/abdominal pain | No=0; Yes=1 |
| 41 | Hip pain | No=0; Yes=1 |
| 42 | Knee pain | No=0; Yes=1 |
| 43 | Whole-body pain | No=0; Yes=1 |
| 44 | Facial pain | No=0; Yes=1 |
| 45 | Sciatica | No=0; Yes=1 |
| 46 | Gastric reflux | No=0; Yes=1 |
| 47 | Hiatus hernia | No=0; Yes=1 |
| 48 | Gall stones | No=0; Yes=1 |
| 49 | Diverticulitis | No=0; Yes=1 |

**Supplementary Table 3** List of the 109 ICD-10 codes and assigned weights for construction of the Hospital Frailty Risk Score

| # | ICD-10 code | Description | Weight |
|---|---|---|---|
| 1 | F00 | Dementia in Alzheimer's disease | 7.1 |
| 2 | G81 | Hemiplegia | 4.4 |
| 3 | G30 | Alzheimer's disease | 4.0 |
| 4 | I69 | Sequelae of cerebrovascular disease (secondary codes) | 3.7 |
| 5 | R29 | Other symptoms and signs involving the nervous and musculoskeletal systems (R29.6 Tendency to fall) | 3.6 |
| 6 | N39 | Other disorders of urinary system (includes urinary tract infection and urinary incontinence) | 3.2 |
| 7 | F05 | Delirium, not induced by alcohol and other psychoactive substances | 3.2 |
| 8 | W19 | Unspecified fall | 3.2 |
| 9 | S00 | Superficial injury of head | 3.2 |
| 10 | R31 | Unspecified hematuria | 3.0 |
| 11 | B96 | Other bacterial agents as the cause of diseases classified to other chapters (secondary code) | 2.9 |
| 12 | R41 | Other symptoms and signs involving cognitive functions and awareness | 2.7 |
| 13 | R26 | Abnormalities of gait and mobility | 2.6 |
| 14 | I67 | Other cerebrovascular diseases | 2.6 |
| 15 | R56 | Convulsions, not elsewhere classified | 2.6 |
| 16 | R40 | Somnolence, stupor and coma | 2.5 |
| 17 | T83 | Complications of genitourinary prosthetic devices, implants and grafts | 2.4 |
| 18 | S06 | Intracranial injury | 2.4 |
| 19 | S42 | Fracture of shoulder and upper arm | 2.3 |
| 20 | E87 | Other disorders of fluid, electrolyte and acid- base balance | 2.3 |
| 21 | M25 | Other joint disorders, not elsewhere classified | 2.3 |
| 22 | E86 | Volume depletion | 2.3 |
| 23 | R54 | Senility | 2.2 |
| 24 | Z50 | Care involving use of rehabilitation procedures | 2.1 |
| 25 | F03 | Unspecified dementia | 2.1 |
| 26 | W18 | Other fall on same level | 2.1 |
| 27 | Z75 | Problems related to medical facilities and other health care | 2.0 |
| 28 | F01 | Vascular dementia | 2.0 |
| 29 | S80 | Superficial injury of lower leg | 2.0 |
| 30 | L03 | Cellulitis | 2.0 |
| 31 | H54 | Blindness and low vision | 1.9 |
| 32 | E53 | Deficiency of other B group vitamins | 1.9 |
| 33 | Z60 | Problems related to social environment | 1.8 |
| 34 | G20 | Parkinson's disease | 1.8 |
| 35 | R55 | Syncope and collapse | 1.8 |
| 36 | S22 | Fracture of rib(s), sternum and thoracic spine | 1.8 |
| 37 | K59 | Other functional intestinal disorders | 1.8 |
| 38 | N17 | Acute renal failure | 1.8 |
| 39 | L89 | Decubitus ulcer | 1.7 |
| 40 | Z22 | Carrier of infectious disease | 1.7 |
| 41 | B95 | Streptococcus and staphylococcus as the cause of diseases classified to other chapters | 1.7 |
| 42 | L97 | Ulcer of lower limb, not elsewhere classified | 1.6 |
| 43 | R44 | Other symptoms and signs involving general sensations and perceptions | 1.6 |
| 44 | K26 | Duodenal ulcer | 1.6 |
| 45 | I95 | Hypotension | 1.6 |
| 46 | N19 | Unspecified renal failure | 1.6 |
| 47 | A41 | Other septicemia | 1.6 |
| 48 | Z87 | Personal history of other diseases and conditions | 1.5 |
| 49 | J96 | Respiratory failure, not elsewhere classified | 1.5 |
| 50 | X59 | Exposure to unspecified factor | 1.5 |
| 51 | M19 | Other arthrosis | 1.5 |
| 52 | G40 | Epilepsy | 1.5 |
| 53 | M81 | Osteoporosis without pathological fracture | 1.4 |

**Supplementary Table 3** (*continued*)

| # | ICD-10 code | Description | Weight |
|---|---|---|---|
| 54 | S72 | Fracture of femur | 1.4 |
| 55 | S32 | Fracture of lumbar spine and pelvis | 1.4 |
| 56 | E16 | Other disorders of pancreatic internal secretion | 1.4 |
| 57 | R94 | Abnormal results of function studies | 1.4 |
| 58 | N18 | Chronic renal failure | 1.4 |
| 59 | R33 | Retention of urine | 1.3 |
| 60 | R69 | Unknown and unspecified causes of morbidity | 1.3 |
| 61 | N28 | Other disorders of kidney and ureter, not elsewhere classified | 1.3 |
| 62 | R32 | Unspecified urinary incontinence | 1.2 |
| 63 | G31 | Other degenerative diseases of nervous system, not elsewhere classified | 1.2 |
| 64 | Y95 | Nosocomial condition | 1.2 |
| 65 | S09 | Other and unspecified injuries of head | 1.2 |
| 66 | R45 | Symptoms and signs involving emotional state | 1.2 |
| 67 | G45 | Transient cerebral ischemic attacks and related syndromes | 1.2 |
| 68 | Z74 | Problems related to care-provider dependency | 1.1 |
| 69 | M79 | Other soft tissue disorders, not elsewhere classified | 1.1 |
| 70 | W06 | Fall involving bed | 1.1 |
| 71 | S01 | Open wound of head | 1.1 |
| 72 | A04 | Other bacterial intestinal infections | 1.1 |
| 73 | A09 | Diarrhea and gastroenteritis of presumed infectious origin | 1.1 |
| 74 | J18 | Pneumonia, organism unspecified | 1.1 |
| 75 | J69 | Pneumonitis due to solids and liquids | 1.0 |
| 76 | R47 | Speech disturbances, not elsewhere classified | 1.0 |
| 77 | E55 | Vitamin D deficiency | 1.0 |
| 78 | Z93 | Artificial opening status | 1.0 |
| 79 | R02 | Gangrene, not elsewhere classified | 1.0 |
| 80 | R63 | Symptoms and signs concerning food and fluid intake | 0.9 |
| 81 | H91 | Other hearing loss | 0.9 |
| 82 | W10 | Fall on and from stairs and steps | 0.9 |
| 83 | W01 | Fall on same level from slipping, tripping and stumbling | 0.9 |
| 84 | E05 | Thyrotoxicosis [hyperthyroidism] | 0.9 |
| 85 | M41 | Scoliosis | 0.9 |
| 86 | R13 | Dysphagia | 0.8 |
| 87 | Z99 | Dependence on enabling machines and devices | 0.8 |
| 88 | U80 | Agent resistant to penicillin and related antibiotics | 0.8 |
| 89 | M80 | Osteoporosis with pathological fracture | 0.8 |
| 90 | K92 | Other diseases of digestive system | 0.8 |
| 91 | I63 | Cerebral Infarction | 0.8 |
| 92 | N20 | Calculus of kidney and ureter | 0.7 |
| 93 | F10 | Mental and behavioral disorders due to use of alcohol | 0.7 |
| 94 | Y84 | Other medical procedures as the cause of abnormal reaction of the patient | 0.7 |
| 95 | R00 | Abnormalities of heartbeat | 0.7 |
| 96 | J22 | Unspecified acute lower respiratory infection | 0.7 |
| 97 | Z73 | Problems related to life-management difficulty | 0.6 |
| 98 | R79 | Other abnormal findings of blood chemistry | 0.6 |
| 99 | Z91 | Personal history of risk-factors, not elsewhere classified | 0.5 |
| 100 | S51 | Open wound of forearm | 0.5 |
| 101 | F32 | Depressive episode | 0.5 |
| 102 | M48 | Spinal stenosis (secondary code only) | 0.5 |
| 103 | E83 | Disorders of mineral metabolism | 0.4 |
| 104 | M15 | Polyarthritis | 0.4 |
| 105 | D64 | Other anemias | 0.4 |
| 106 | L08 | Other local infections of skin and subcutaneous tissue | 0.4 |
| 107 | R11 | Nausea and vomiting | 0.3 |
| 108 | K52 | Other noninfective gastroenteritis and colitis | 0.3 |
| 109 | R50 | Fever of unknown origin | 0.1 |

Abbreviation: *ICD-10*, International Statistical Classification of Diseases and Related Health Problems, Tenth Revision

**Supplementary Table 4** List of the 17 comorbidities, assigned weights and associated ICD-10 codes for construction of Charlson Comorbidity Index

| # | Condition | Weight | ICD-10 code |
|---|---|---|---|
| 1 | Myocardial infarction | 1 | I21, I22, I25.2 |
| 2 | Congestive heart failure | 1 | I50 |
| 3 | Peripheral vascular disease | 1 | I71, I73.9, I79.0, R02, Z95.8, Z95.9 |
| 4 | Cerebral vascular disease | 1 | G45, G46, I60–I64, I67–I69 |
| 5 | Dementia | 1 | F00–F02, F05.1 |
| 6 | Pulmonary disease | 1 | J40–J47, J60–J67 |
| 7 | Connective tissue disorder | 1 | M05, M06, M32–M34, M35.3 |
| 8 | Peptic ulcer | 1 | K25–K28 |
| 9 | Liver disease | 1 | K70.2, K70.3, K71.7, K73, K74 |
| 10 | Diabetes | 1 | E10–E14, excluding subdivisions 2–5 |
| 11 | Diabetes complications | 2 | E10–E14, subdivisions 2–5 |
| 12 | Paraplegia | 2 | G04.1, G81, G82 |
| 13 | Renal disease | 2 | N01, N03, N18, N19, N25, N05.2–N05.6, N07.2–N07.4, |
| 14 | Cancer | 2 | C00–C41, C43, C45–C76, C81–C85, C88, C90–C97 |
| 15 | Metastatic cancer | 3 | C77–C80 |
| 16 | Severe liver disease | 3 | K72.1, K72.9, K76.6, K76.7 |
| 17 | AIDS | 6 | B20–B24 |

Abbreviations: *AIDS*, Acquired immune deficiency syndrome; *ICD-10*, International Statistical Classification of Diseases and Related Health Problems, Tenth Revision

**Supplementary Table 5** Characteristics of individuals with and without hospital data

| Characteristic | With hospital data (n=370,660) | Without hospital data[a] (n=58,094) | $p$[b] |
|---|---|---|---|
| Deaths, n (%) | 2,291 (0.6) | 60 (0.1) | <0.001 |
| Age, mean (SD), years | 68.5 (8.1) | 65.6 (7.7) | <0.001 |
| Age category, n (%) | | | <0.001 |
| <65 | 119,127 (32.1) | 26,173 (45.1) | |
| 65–74 | 149,822 (40.4) | 23,685 (40.8) | |
| ≥75 | 101,711 (27.4) | 8,236 (14.2) | |
| Sex, n (%) | | | <0.001 |
| Female | 207,044 (55.9) | 29,600 (51.0) | |
| Male | 163,616 (44.1) | 28,494 (49.0) | |
| Ethnicity, n (%) | | | <0.001 |
| White | 349,300 (95.6) | 54,110 (93.5) | |
| Asian | 8,313 (2.3) | 1,593 (2.8) | |
| Black | 6,229 (1.7) | 1,122 (1.9) | |
| Others | 5,576 (1.5) | 1,065 (1.8) | |
| Smoking status, n (%) | | | <0.001 |
| Never | 201,756 (54.6) | 35,951 (62.0) | |
| Previous | 130,554 (35.4) | 16,923 (29.2) | |
| Current | 37,038 (10.0) | 5,073 (8.8) | |
| Education, n (%) | | | <0.001 |
| Low | 63,227 (17.2) | 5,404 (9.4) | |
| Intermediate | 187,950 (51.2) | 28,385 (49.2) | |
| High | 115,753 (31.6) | 23,868 (41.4) | |
| Income, n (%) | | | <0.001 |
| <£18,000 | 72,821 (23.0) | 7,096 (13.9) | |
| £18,000–30,999 | 82,596 (26.1) | 10,807 (21.1) | |
| £31,000–51,999 | 83,099 (26.3) | 14,304 (28.0) | |
| ≥£52,000 | 77,998 (24.6) | 18,964 (37.1) | |
| Townsend deprivation quintile, n (%) | | | <0.001 |
| 1 (least deprived) | 74,039 (20.0) | 12,405 (21.4) | |
| 2 | 75,329 (20.4) | 12,079 (20.8) | |
| 3 | 75,128 (20.3) | 11,677 (20.1) | |
| 4 | 74,216 (20.1) | 11,596 (20.0) | |
| 5 (most deprived) | 71,511 (19.3) | 10,262 (17.7) | |
| Frailty phenotype category, n (%) | | | <0.001 |
| Non-frail | 209,784 (56.6) | 39,036 (67.2) | |
| Pre-frail | 146,773 (39.6) | 18,419 (31.7) | |
| Frail | 14,103 (3.8) | 639 (1.1) | |
| Frailty index category, n (%) | | | <0.001 |
| Relatively fit | 18,956 (5.1) | 6,942 (12.0) | |
| Less fit | 132,051 (35.6) | 30,098 (51.8) | |
| Least fit | 170,396 (46.0) | 19,340 (33.3) | |
| Frail | 49,257 (13.3) | 1,714 (3.0) | |

[a] Resided in England at baseline and without hospital data between 1980 and 2020
[b] Based on t-tests or Mann–Whitney *U* tests for continuous variables and $\chi^2$-tests for categorical variables
Abbreviations: *IQR*, interquartile range; *SD*, standard deviation

**Supplementary Table 6** Characteristics of the COVID-19 positive subsample (n=2,287) by the source of the disease status

| | Confirmed COVID-19 cases (n=2,201; deaths=383)[a] | Died of COVID-19 without positive test record (n=86)[b] | $p$[c] |
|---|---|---|---|
| Age (year), mean (SD) | 67.9 (9.1) | 75.4 (6.0) | <0.001 |
| Age category, n (%) | | | <0.001 |
| <65 | 857 (38.9) | 6 (7.0) | |
| 65–74 | 645 (29.3) | 20 (23.3) | |
| ≥75 | 699 (31.8) | 60 (69.8) | |
| Sex, n (%) | | | 0.17 |
| Female | 1,062 (48.3) | 35 (40.7) | |
| Male | 1,139 (51.8) | 51 (59.3) | |
| Ethnicity, n (%) | | | 0.20 |
| White | 1,925 (87.8) | 82 (95.4) | |
| Asian | 110 (5.0) | 2 (2.3) | |
| Black | 103 (4.7) | 1 (1.2) | |
| Others | 55 (2.5) | 1 (1.2) | |
| Smoking status, n (%) | | | 0.23 |
| Never | 1,070 (49.0) | 34 (39.5) | |
| Previous | 847 (38.8) | 40 (46.5) | |
| Current | 268 (12.3) | 12 (14.0) | |
| Education, n (%) | | | 0.76 |
| Low | 530 (24.4) | 23 (27.7) | |
| Intermediate | 1,097 (50.6) | 39 (47.0) | |
| High | 543 (25.0) | 21 (25.3) | |
| Income, n (%) | | | 0.015 |
| <£18,000 | 580 (31.5) | 31 (40.8) | |
| £18,000–30,999 | 459 (24.9) | 25 (32.9) | |
| £31,000–51,999 | 417 (22.7) | 14 (18.4) | |
| ≥£52,000 | 385 (20.9) | 6 (7.9) | |
| Townsend deprivation quintile, n (%) | | | 0.36 |
| 1 (least deprived) | 329 (15.0) | 14 (16.3) | |
| 2 | 366 (16.6) | 13 (15.1) | |
| 3 | 394 (17.9) | 22 (25.6) | |
| 4 | 465 (21.1) | 18 (20.9) | |
| 5 (most deprived) | 646 (29.4) | 19 (22.1) | |
| Frailty phenotype category, n (%) | | | 0.07 |
| Non-frail | 1,097 (49.8) | 34 (39.5) | |
| Pre-frail | 925 (42.0) | 47 (54.7) | |
| Frail | 179 (8.1) | 5 (5.8) | |
| Frailty index category, n (%) | | | 0.30 |
| Relatively fit | 96 (4.4) | 2 (2.3) | |
| Less fit | 678 (30.8) | 20 (23.3) | |
| Least fit | 996 (45.3) | 46 (53.5) | |
| Frail | 431 (19.6) | 18 (20.9) | |
| Hospital Frailty Risk Score category, n (%) | | | 0.001 |
| Low risk | 1,730 (78.6) | 55 (64.0) | |
| Intermediate risk | 220 (10.0) | 10 (11.6) | |
| High risk | 251 (11.4) | 21 (24.4) | |
| Charlson Comorbidity Index score, mean (SD) | 0.93 (1.73) | 1.09 (1.64) | 0.16 |

[a] Positive in at least one of the COVID-19 tests or recorded as a COVID-19 inpatient in hospital data
[b] With COVID-19 as the primary or contributory cause of death but were not recorded as tested positive or was a COVID-19 inpatient
[c] Based on t-tests or Mann–Whitney *U* tests for continuous variables and $\chi^2$-tests for categorical variables
Abbreviations: *IQR*, interquartile range; *SD*, standard deviation

**Supplementary Table 7** Spearman's rank correlation coefficients among the frailty measures and comorbidity

| Variable | FP score | FI score | HFRS score | CCI score |
|---|---|---|---|---|
| FP score[a] | 1 | | | |
| FI score | 0.341* | 1 | | |
| HFRS score | 0.115* | 0.189* | 1 | |
| CCI score | 0.123* | 0.216* | 0.554* | 1 |

[a] FP is considered as a continuous variable using the score across the five criteria (ranging from 0 to 5)
* $p$ <0.001
Abbreviations: *CCI*, Charlson Comorbidity Index; *FI*, frailty index; *FP*, frailty phenotype; *HFRS*, hospital frailty risk score

**Supplementary Table 8** Associations between different frailty and comorbidity measures, and being COVID-19 positive (n=428,754)

| Variable | Model 1[a] | Model 2[a] | Model 3[a] | Model 4[a] | Model 5[a] | Model 6[a] |
|---|---|---|---|---|---|---|
| | OR (95% CI) | OR (95% CI) | OR (95% CI) | OR (95% CI) | OR (95% CI) | OR (95% CI) |
| Age | 1.00 (1.00–1.01) | 1.00 (0.99–1.00) | 1.00 (0.99–1.00) | 0.98 (0.98–0.99)* | 0.99 (0.98–0.99)* | 0.98 (0.98–0.99)* |
| Male sex | 1.34 (1.23–1.45)* | 1.38 (1.27–1.50)* | 1.38 (1.27–1.50)* | 1.30 (1.19–1.41)* | 1.26 (1.16–1.36)* | 1.30 (1.20–1.41)* |
| FP category (ref. non-frail) | | | | | | |
| Pre-frail | - | 1.32 (1.21–1.44)* | - | - | - | 1.11 (1.01–1.21)* |
| Frail | - | 2.89 (2.47–3.38)* | - | - | - | 1.48 (1.24–1.77)* |
| FI category (ref. relatively fit) | | | | | | |
| Less fit | - | - | 1.17 (0.94–1.44) | - | - | 1.14 (0.92–1.41) |
| Least fit | - | - | 1.51 (1.23–1.86)* | - | - | 1.30 (1.06–1.61)* |
| Frail | - | - | 2.48 (1.99-3.09)* | - | - | 1.44 (1.14–1.81)* |
| HFRS category (ref. low risk) | | | | | | |
| Intermediate risk | - | - | - | 3.66 (3.18–4.21)* | - | 2.55 (2.18–2.97)* |
| High risk | - | - | - | 19.29 (16.82–22.12)* | - | 10.72 (9.02–12.72)* |
| CCI score | - | - | - | - | 1.45 (1.42–1.49)* | 1.17 (1.13–1.21)* |
| AUC (95% CI) | 0.54 (0.53–0.55) | 0.57 (0.56–0.58) | 0.58 (0.57–0.59) | 0.64 (0.62–0.65) | 0.64 (0.63–0.65) | 0.67 (0.66–0.68) |

[a] Model 1: age (continuous) and sex; models 2–5: added FP, FI, HFRS, and CCI as independent variables respectively; model 6: all listed variables were mutually adjusted for
* Significant with a false discovery rate corrected significance level at 0.041
Abbreviations: *AUC*, area under the receiver operating characteristic curves; *CCI*, Charlson Comorbidity Index; *CI*, confidence interval; *FI*, frailty index; *FP*, frailty phenotype; *HFRS*, Hospital Frailty Risk Score; *OR*, odds ratio.

**Supplementary Table 9** Associations between the Hospital Frailty Risk Score and COVID-19 mortality stratified by age in the full sample and COVID-19 positive subsample

| Variable | Age category | | | $P_{interaction}$[a] |
|---|---|---|---|---|
| | <65 years | 65–74 years | ≥75 years | |
| | OR (95% CI) | OR (95% CI) | OR (95% CI) | |
| **(a) Full sample (n=428,754)** | | | | |
| HFRS category (ref. low risk) | | | | |
| Intermediate risk | 10.60 (3.66–30.68)* | 6.53 (3.90–10.93)* | 4.70 (3.38–6.54)* | 0.003 |
| High risk | 77.71 (26.68–226.29)* | 34.19 (20.59–56.76)* | 18.71 (13.78–25.41)* | |
| Male sex | 1.14 (0.56–2.31) | 1.89 (1.29–2.76)* | 2.13 (1.63–2.78)* | |
| AUC (95% CI) | 0.62 (0.51–0.72) | 0.71 (0.65–0.76) | 0.73 (0.69–0.76) | |
| **(b) COVID-19 positive sample[b] (n=2,287)** | | | | |
| HFRS category (ref. low risk) | | | | |
| Intermediate risk | 5.27 (1.67–16.62)* | 2.23 (1.23–4.06)* | 1.28 (0.85–1.93) | <0.001 |
| High risk | 15.39 (4.41–53.64)* | 1.70 (0.96–3.00) | 1.15 (0.79–1.67) | |
| Male sex | 1.04 (0.49–2.19) | 1.54 (1.01–2.34) | 1.54 (1.12–2.12) | |
| AUC (95% CI) | 0.61 (0.50–0.72) | 0.60 (0.54–0.65) | 0.57 (0.52–0.61) | |

[a] $P_{interaction}$ represents the *p*-values of the interaction terms fitted between the HFRS and age as continuous variables
[b] The COVID-19 positive subsample is nested within the full sample
* Significant with a false discovery rate corrected significance level at 0.033
Abbreviations: *AUC*, area under the receiver operating characteristic curves; *CI*, confidence interval; *HFRS*, Hospital Frailty Risk Score; *OR*, odds ratio.

**Supplementary Table 10** Associations between different frailty and comorbidity measures, and mortality due to COVID-19 and other causes in the full sample and COVID-19 positive subsample

| Variable | Model 1[a] | | Model 2[a] | | Model 3[a] | | Model 4[a] | | Model 5[a] | |
|---|---|---|---|---|---|---|---|---|---|---|
| | COVID-19 mortality | Other deaths[b] | COVID-19 mortality | Other deaths[b] | COVID-19 mortality | Other deaths[b] | COVID-19 mortality | Other deaths[b] | COVID-19 mortality | Other deaths[b] |
| | OR (95% CI) | OR (95% CI) | OR (95% CI) | OR (95% CI) | OR (95% CI) | OR (95% CI) | OR (95% CI) | OR (95% CI) | OR (95% CI) | OR (95% CI) |
| **(a) Full sample (n=428,754)** | | | | | | | | | | |
| Age | 1.14 (1.12–1.16)* | 1.12 (1.11–1.13)* | 1.14 (1.12–1.16)* | 1.12 (1.11–1.13)* | 1.11 (1.09–1.13)* | 1.09 (1.08–1.10)* | 1.12 (1.10–1.14)* | 1.09 (1.08–1.10)* | 1.11 (1.09–1.13)* | 1.08 (1.07–1.09)* |
| Male sex | 2.16 (1.76–2.66)* | 1.62 (1.48–1.78)* | 2.15 (1.75–2.65)* | 1.62 (1.48–1.77)* | 1.98 (1.61–2.44)* | 1.51 (1.38–1.65)* | 1.80 (1.46–2.21)* | 1.26 (1.15–1.38)* | 1.93 (1.57–2.38)* | 1.26 (1.14–1.38)* |
| FP category (ref. non-frail) | | | | | | | | | | |
| Pre-frail | 1.55 (1.26–1.91)* | 1.34 (1.22–1.47)* | - | - | - | - | - | - | 1.19 (0.95–1.48) | 1.01 (0.91–1.11) |
| Frail | 3.82 (2.69–5.41)* | 2.94 (2.49–3.47)* | - | - | - | - | - | - | 1.54 (1.04–2.28) | 1.10 (0.90–1.34) |
| FI category (ref. relatively fit) | | | | | | | | | | |
| Less fit | - | - | 1.41 (0.74–2.71) | 1.10 (0.86–1.41) | - | - | - | - | 1.32 (0.69–2.54) | 0.99 (0.77–1.27) |
| Least fit | - | - | 2.00 (1.06–3.79) | 1.40 (1.09–1.78)* | - | - | - | - | 1.49 (0.78–2.84) | 0.91 (0.71–1.16) |
| Frail | - | - | 3.75 (1.05–7.21)* | 2.36 (1.83–3.04)* | - | - | - | - | 1.63 (0.83–3.19) | 0.75 (0.57–0.98) |
| HFRS category (ref. low risk) | | | | | | | | | | |
| Intermediate risk | - | - | - | - | 5.42 (4.13–7.11)* | 8.47 (7.58–9.46)* | - | - | 3.22 (2.38–4.35)* | 2.91 (2.55–3.32)* |
| High risk | - | - | - | - | 24.57 (18.94–31.87)* | 25.47 (22.43–28.92)* | - | - | 11.41 (8.27–15.74)* | 5.58 (4.77–6.53)* |
| CCI score | - | - | - | - | - | - | 1.59 (1.52–1.66)* | 1.86 (1.82–1.89)* | 1.26 (1.19–1.34)* | 1.64 (1.60–1.68)* |

**Supplementary Table 10** (*continued*)

| Variable | Model 1[a] | | Model 2[a] | | Model 3[a] | | Model 4[a] | | Model 5[a] | |
|---|---|---|---|---|---|---|---|---|---|---|
| | COVID-19 mortality | Other deaths[b] | COVID-19 mortality | Other deaths[b] | COVID-19 mortality | Other deaths[b] | COVID-19 mortality | Other deaths[b] | COVID-19 mortality | Other deaths[b] |
| | OR (95% CI) | OR (95% CI) | OR (95% CI) | OR (95% CI) | OR (95% CI) | OR (95% CI) | OR (95% CI) | OR (95% CI) | OR (95% CI) | OR (95% CI) |
| **(b) COVID-19 positive subsample[c] (n=2,287)** | | | | | | | | | | |
| Age | 1.13 (1.11–1.15)* | 1.10 (1.07–1.14)* | 1.13 (1.11–1.15)* | 1.10 (1.06–1.14)* | 1.13 (1.11–1.15)* | 1.08 (1.05–1.12)* | 1.13 (1.11–1.15)* | 1.08 (1.05–1.12)* | 1.13 (1.11–1.15)* | 1.09 (1.05–1.13)* |
| Male sex | 1.55 (1.22–1.98)* | 1.74 (1.07–2.83)* | 1.55 (1.22–1.97)* | 1.77 (1.09–2.87)* | 1.54 (1.21–1.96)* | 1.74 (1.07–2.82)* | 1.47 (1.16–1.88)* | 1.53 (0.93–2.50) | 1.50 (1.17–1.91)* | 1.52 (0.92–2.51) |
| FP category (ref. non-frail) | | | | | | | | | | |
| Pre-frail | 1.19 (0.93–1.52) | 1.15 (0.70–1.87) | - | - | - | - | - | - | 1.11 (0.85–1.44) | 0.96 (0.57–1.63) |
| Frail | 1.29 (0.85–1.96) | 1.53 (0.71–3.31) | - | - | - | - | - | - | 1.03 (0.65–1.63) | 0.84 (0.34–2.07) |
| FI category (ref. relatively fit) | | | | | | | | | | |
| Less fit | - | - | 1.07 (0.51–2.24) | 0.45 (0.14–1.41) | - | - | - | - | 1.04 (0.49–2.17) | 0.40 (0.12–1.27) |
| Least fit | - | - | 1.19 (0.58–2.45) | 0.65 (0.22–1.91) | - | - | - | - | 1.08 (0.52–2.24) | 0.52 (0.17–1.56) |
| Frail | - | - | 1.35 (0.65–2.84) | 0.82 (0.27–2.52) | - | - | - | - | 1.05 (0.49–2.24) | 0.45 (0.14–1.50) |
| HFRS category (ref. low risk) | | | | | | | | | | |
| Intermediate risk | - | - | - | - | 1.79 (1.28–2.52)* | 3.07 (1.68–5.60)* | - | - | 1.29 (0.89–1.86) | 1.36 (0.70–2.65) |
| High risk | - | - | - | - | 1.41 (1.03–1.93) | 2.23 (1.23–4.05)* | - | - | 0.89 (0.61–1.29) | 0.71 (0.34–1.45) |
| CCI score | - | - | - | - | - | - | 1.20 (1.12–1.27)* | 1.44 (1.32–1.58)* | 1.19 (1.11–1.28)* | 1.47 (1.32–1.65)* |

[a] Associations were assessed using multinomial logistic regression models, in which "COVID-19 mortality" and "other deaths" were compared to those who were alive as of. Model 1–4: FP, FI, HFRS, and CCI as independent variables respectively, adjusted for age and sex; model 6: all listed variables were mutually adjusted for

[b] Other deaths are individuals died between March 1 and September 21, 2020 whose primary and contributory causes of death do not include COVID-19

[c] The COVID-19 positive subsample is nested within the full sample

* Significant with a false discovery rate corrected significance level at 0.031

Abbreviations: *CCI*, Charlson Comorbidity Index; *CI*, confidence interval; *FI*, frailty index; *FP*, frailty phenotype; *HFRS*, Hospital Frailty Risk Score; *OR*, odds ratio.
